# Cluster-randomized trial of digital adherence technologies and differentiated care to reduce poor end-of-treatment outcomes and recurrence among adults with drug-sensitive pulmonary TB in Ethiopia

**DOI:** 10.1101/2024.05.09.24307117

**Authors:** Amare W Tadesse, Mamush Sahile, Nicola Foster, Christopher Finn McQuaid, Gedion Teferra Weldemichael, Tofik Abdurhman, Zemedu Mohammed, Mahilet Belachew, Amanuel Shiferaw, Demelash Assefa, Demekech Gadissa, Hiwot Yazew, Nuria Yakob, Zewdneh Shewamene, Lara Goscé, Job van Rest, Norma Madden, Salome Charalambous, Kristian van Kalmthout, Ahmed Bedru, Taye Letta, Degu Jerene, Katherine L Fielding

## Abstract

**Background:** The impact of Digital Adherence Technologies (DATs) on long-term tuberculosis treatment outcomes remains unclear. We aimed to assess the effectiveness of DATs and differentiated care in improving tuberculosis treatment outcomes and recurrence.

**Methods:** We conducted a pragmatic cluster-randomised trial in Ethiopia. Seventy-eight health facilities (clusters) were randomised to three arms (1:1:1): smart pillbox, medication labels, or standard of care. Adults (≥18 years) with drug-sensitive pulmonary tuberculosis on a fixed-dose combination tuberculosis treatment regimen were enrolled and followed-up for 12 months after treatment initiation. Those in the pillbox arm received a pillbox with customisable audio-visual reminders, while participants in the label arm received their TB medication with a weekly unique code label. Opening the box or texting the code prompted real-time dose logging on the adherence platform, facilitating differentiated response by a healthcare worker. The primary outcome comprised death, loss to follow-up, treatment failure, switch to drug-resistant tuberculosis treatment, or recurrence; secondary outcomes included loss to follow-up. Analysis accounted for clustered design with multiple imputation for the primary outcome. The trial is complete and registered with PACTR202008776694999.

**Findings:** From 24/05/2021-08/08/2022, 8477 individuals undergoing tuberculosis treatment were assessed for eligibility, and 3885 participants enrolled, of whom 3858 were included in the intention-to-treat population. The median age was 30 years and 41% were female. At 12 months, using multiple imputation, neither the pillbox (adjusted OR 1.04, 95% CI: 0.74-1.45; adjusted risk difference, 1.0 percentage points, 95% CI -1.2 to 3.1) nor the label (adjusted OR 1.14, 95%CI: 0.83-1.61; adjusted risk difference, 0.4 percentage points, 95% CI -1.8 to 2.6) interventions reduced the risk of the primary composite outcome. Results were similar in complete case and per-protocol analyses.

**Interpretation:** The DAT interventions showed no reduction in unfavourable outcomes. This emphasizes the necessity to optimise DATs to enhance TB management strategies and treatment outcomes.

## Introduction

Tuberculosis (TB) remains a significant global health challenge and the leading cause of mortality from a single infectious agent, except during the coronavirus (COVID-19) pandemic in 2022 with an estimated 10.6 million individuals developing TB disease and 1.30 million deaths worldwide (1). Although global data indicate some improvement in treatment outcomes for people treated for TB, the emergence of drug-resistant TB, gaps in detection and reporting of TB cases, and poor treatment outcomes persist as challenges in global control efforts (1).

Treatment for drug-sensitive (DS) pulmonary TB involves taking daily treatment for 6 months, 2 months of four drugs, and 4 months of two drugs, primarily using a fixed-dose combination. Treatment adherence is critical for achieving a cure, averting transmission, and reducing the recurrence of tuberculosis (2, 3). Directly Observed Treatment (DOT) has been a globally recommended strategy for TB treatment supervision since 1994 (4). The success of DOT relies on direct observation of people with TB taking their medication, where the need for consistent and reliable supervision during treatment administration places a significant burden on both the individual with TB and the health system (5-7). Innovative person-centred solutions, such as the use of digital adherence technologies (DATs), to enhance treatment supervision and monitoring, have emerged as alternative strategies to support treatment adherence (8, 9). Such technologies are used in other disease areas (10), and increasingly used during COVID-19 when in-person care was challenging. WHO conditionally recommends the use of digital tools, including SMS-based reminders, video-supported therapy, and smart pillboxes, for TB care and support (11).

Recent DAT trials have reported mixed results on the effectiveness of DATs in improving TB treatment and health outcomes (12), including large-scale trials (13-15). Many trials have shown an impact on treatment adherence (13, 16-23) but few have demonstrated an impact on improving end of treatment outcomes(16, 24, 25). Including TB recurrence in the endpoint definition, as is standard for DS-TB treatment trials, provides a more sensitive measure of the success of the intervention, though few DAT trials have attempted to do so (13, 14).

Ethiopia is one of the 30 high TB burden countries globally (1). Although in 2021 compared with 2015, there had been a 20% reduction in TB incidence and a 34% reduction in mortality (26), these gains have been challenged by the impacts of the COVID-19 pandemic on TB care services and the health system (1). As part of The Adherence Support Coalition to End TB (ASCENT) project (27), we conducted a three-arm pragmatic cluster-randomized trial in Ethiopia to assess the effectiveness of smart pillboxes and medication labels integrated with an adherence data platform among people on treatment for DS pulmonary TB. These interventions aimed to enable daily monitoring of adherence data and provide differentiated care responses by healthcare providers, to reduce unfavourable treatment outcomes, including TB recurrence. Given TB care is implemented at the health facility level, cluster randomisation at that level was chosen to reduce contamination between study arms and for logistical convenience.

## Methods

### Study design

We conducted a pragmatic cluster-randomized trial, in two regions of Ethiopia, with the health facility, the cluster, as the unit of randomization. The trial design is reported elsewhere (27). Health facilities were randomised (ratio 1:1:1) to either the (i) smart pillbox or (ii) medication labels, both with daily monitoring and differentiated response to patient adherence, or (iii) standard of care arm. This trial was part of the ASCENT consortium which conducted trials in four other countries, under a separate protocol and reported separately due to a different trial design (two-arm studies with no follow-up for recurrence) (15, 28). The pragmatic trial design was chosen as the interest was in how the DAT interventions would work under routine conditions; design choices, therefore, included broad eligibility criteria, delivery of the intervention (primarily implemented by health facility staff), and outcomes relevant to participants. The trial received ethics approval from the Institutional Review Board of the Public Emergency and Health Research Directorate at the Health Bureau of Addis Ababa City Administration and Oromia Region of Ethiopia, the Ethics Committee of the London School of Hygiene & Tropical Medicine, and the WHO Ethical Review Committee.

### Clusters and participants

The clusters were chosen in collaboration with the Ethiopian National TB Programme (NTP) from two regions (Addis Ababa and Oromia) and included small and large, rural and urban, public health facilities. Inclusion criteria were based on having at least 30 notified people with DS-TB in 2018 and being willing to participate in the trial.

People with TB were eligible for the trial if aged ≥18 years with pulmonary DS-TB, started on TB treatment using a fixed-dose combination, and were likely to be in the study area for 12 months. Those critically ill, receiving in-patient care or palliative care were excluded. Written informed consent was obtained from participants enrolled in the trial by the TB Care Provider (TB focal) in each facility.

### Randomisation and masking

Randomisation of clusters to a study arm in a ratio 1:1:1 was conducted by the trial statistician in Stata version 16 using stratification and restriction to help ensure reasonable balance by study arm and stratification to reduce intra-cluster correlation (29). Clusters were stratified into four groups using region and percentage with poor treatment outcomes in 2017–2019 (low/high). Restriction, conducted using TB notifications and outcomes from 2017–2019, was based on the percentage of poor treatment outcomes, number of DS-TB registrations, HIV prevalence among DS-TB, and urban/rural areas.

### Intervention arms

The intervention arms included a DAT for the participant, a web-based platform for adherence monitoring accessed by the TB focal, and a differentiated response to patient adherence, depending on participant DAT engagement, initiated by the TB focal. In addition, all participants had a designated treatment supporter, responsible for ensuring treatment adherence, recording treatment information, identifying adherence barriers, and facilitating retrieval of people with TB who interrupt treatment, as per usual care.

Participants in the pillbox arm received a pillbox to store their medications, which had a daily audio-visual reminder to take treatment at a time in the morning (6:00-11:00 am) pre-arranged with the health care provider. When the pillbox was opened an embedded device sent a signal of its opening to the adherence platform via mobile internet. Participants in the labels arm who had access to a mobile phone (either their own or shared), received their medication with a label attached containing a weekly unique toll-free number. They were expected to send a toll-free text message daily, at the time of taking their medication, which was logged into the platform. Participants with no access to a mobile phone received a pillbox instead. An automated SMS reminder was sent to all participants with access to a mobile phone who did not open their pillbox or send a text (labels arm) by 11:00 am each day.

A dose not recorded by the DAT was assumed to be a proxy for a missed dose. In both arms differentiated response to the DAT engagement data on the platform was initiated by the TB focal person, with escalating activity for higher levels of DAT non-engagement documented. These activities included: a phone call to the patient or their treatment supporter for one or two missed doses; a home visit by a community health worker for persistent missed doses (≥ 5 consecutive doses or multiple times); and a switch to DOT for persistent missed doses of >14 consecutive doses or multiple times missing 7-14 doses). During phone calls with the participant, TB focal ascertained if a dose had been missed, if not the dose report was changed to a manual dose in the platform.

### Standard of care arm

Participants at standard of care facilities received the standard of care in that facility, be it facility or home-based DOT. Briefly pre-COVID-19, the NTP specified this was direct daily observation of medication intake during the intensive phase by a health worker or TB treatment supporter, at a health facility (hospital, health centre or health post), at a patient’s workplace, residence institution, or home. During the continuation phase, the NTP specified medication intake under direct observation by a TB treatment supporter. Missed clinic appointments resulted in a phone call to the individual and missing two consecutive clinic appointments prompted a home visit by a health care worker. These recommendations were likely to be applied variably. During COVID-19, with varying degrees of travel restrictions during participant enrolment and follow-up, the frequency of direct observation changed to weekly or fortnightly in the intensive phase and monthly in the continuation phase. (See supplement for more details).

### Procedures

Following obtaining informed consent, the TB focal person administered a questionnaire collecting information on socio-demographics and household assets. The research team extracted data from the TB register, including HIV status, history of TB, diagnosis of TB, date of treatment start, and the treatment outcome. For participants who were transferred out to a different facility the date of transfer and the treatment outcome and date from the clinic they were transferred to were documented. Twelve months from enrolment, a visit was conducted for all participants who had treatment completion or cure documented in the TB register. A phone call visit was conducted to ascertain, by self-report, any treatment restarts from the end of treatment to the visit. Participants with microbiologically confirmed TB at enrolment were scheduled for a facility visit to collect a research sputum sample (without induction) for culture. Sputum samples were transported to the National Tuberculosis Laboratory in Addis Ababa for using both Löwenstien-Jensen (LJ) and Mycobacterial Growth Indicator Tube (MGIT).

### Outcomes

The primary outcome was the proportion of participants with a composite unfavourable outcome measured over 12 months from treatment start defined as either (i) poor end of treatment outcome of treatment failure, loss to follow-up, death, or multi-drug resistant (MDR) diagnosis/change to MDR treatment regimen >28 days after treatment start; or (ii) TB recurrence between 6-12 months from enrolment among those with treatment outcome of cured or completed treatment. See supplement for more details. The on-treatment outcome was based on TB register data. TB recurrence was defined as being culture positive for Mycobacterium tuberculosis (MTB) on LJ or MGIT from the research sample taken at 12 months or self-report treatment restart at 12 months from treatment start. Secondary outcomes were poor end of treatment outcomes and loss to follow-up during treatment, both were based on TB register data.

### Statistical analysis

The sample size for the primary outcome took into account the clustered design. We used routine data from TB registrations in 2018 in the health facilities to inform the calculations. We assumed the percentage with unfavourable outcome varied between 17-20% in the standard of care arm, a harmonic mean of 50 adults with pulmonary DS-TB per facility over the 12-month enrolment period and the coefficient of variation (k) between 0.25 to 0.35. Assuming 26 facilities per arm and a type I error of 5%, we would have at least 85% power to detect a one-third reduction in the unfavourable outcome with k ≤0.3 and 80% power for k=0.35. In our randomisation, by including the percentage with poor treatment outcomes in our stratification, we hoped to reduce k, and therefore increase power.

The analyses were based on the logistic regression random-effects model, with a random effect to account for clustering at the health facility level. We assumed a fixed effect for the study arm and reported two comparisons; each intervention arm compared with the standard of care. All models adjusted for randomisation strata as a fixed effect. If baseline imbalance for individual and cluster-level covariates was seen, an analysis adjusting for these covariates was conducted. Two populations were used for all outcomes, the intent to treat (ITT) and the per protocol (PP) populations. The ITT population was based on all individuals enrolled, excluding those for whom their TB diagnosis was changed to “not TB” or had an MDR diagnosis or switched to an MDR regimen, or were transferred out within 28 days of treatment start or those previously enrolled in the study. The PP population was intended primarily to capture the intervention effect among those who were exposed to the intervention. In the intervention arms, we therefore excluded those who started the DAT >28 days after treatment initiation or stopped the DAT within the first 112 days of treatment, for reasons other than death or loss to follow-up or poor adherence. Subgroup analyses for the primary outcome and sensitivity analyses were specified a priori. The statistical analysis plan, reviewed by our independent Technical Advisory Group (TAG), detailed all analyses. Multiple imputation for the primary unfavourable outcome in the ITT population was conducted due to loss to follow-up at 12 months, based on 25 imputations.

## Results

From May 24, 2021, to August 08, 2022, 8477 people starting DS-TB treatment (78 health facilities) were screened, of whom 4496 (53.0%) did not fulfil the enrolment criteria, and 96 (1.1%) opted not to give consent to participate, leaving 3885 participants enrolled in the trial. The reasons for not satisfying enrolment criteria were aged < 18 years or extrapulmonary TB (3402), not likely to be in the area for at least 12 months (718), too ill (352), and 20 individuals had incomplete data on eligibility Of the 3885 enrolled, 27 were later excluded due to either an MDR diagnosis in the first 28 days (7), diagnosis changed to not TB (14) or enrolled twice in error (6), leaving 3858 (1258 pillbox, 1305 labels, 1295 standard of care) participants, from 78 health facilities, in the ITT population (Figure 1). Overall, 3.4% of participants (132/3858; 76 from the labels and 56 from the pillbox group) were excluded from the per-protocol population, leaving 3826 participants in this population. The last 12-month follow-up visit occurred on August 16, 2023.

**Figure 1:**
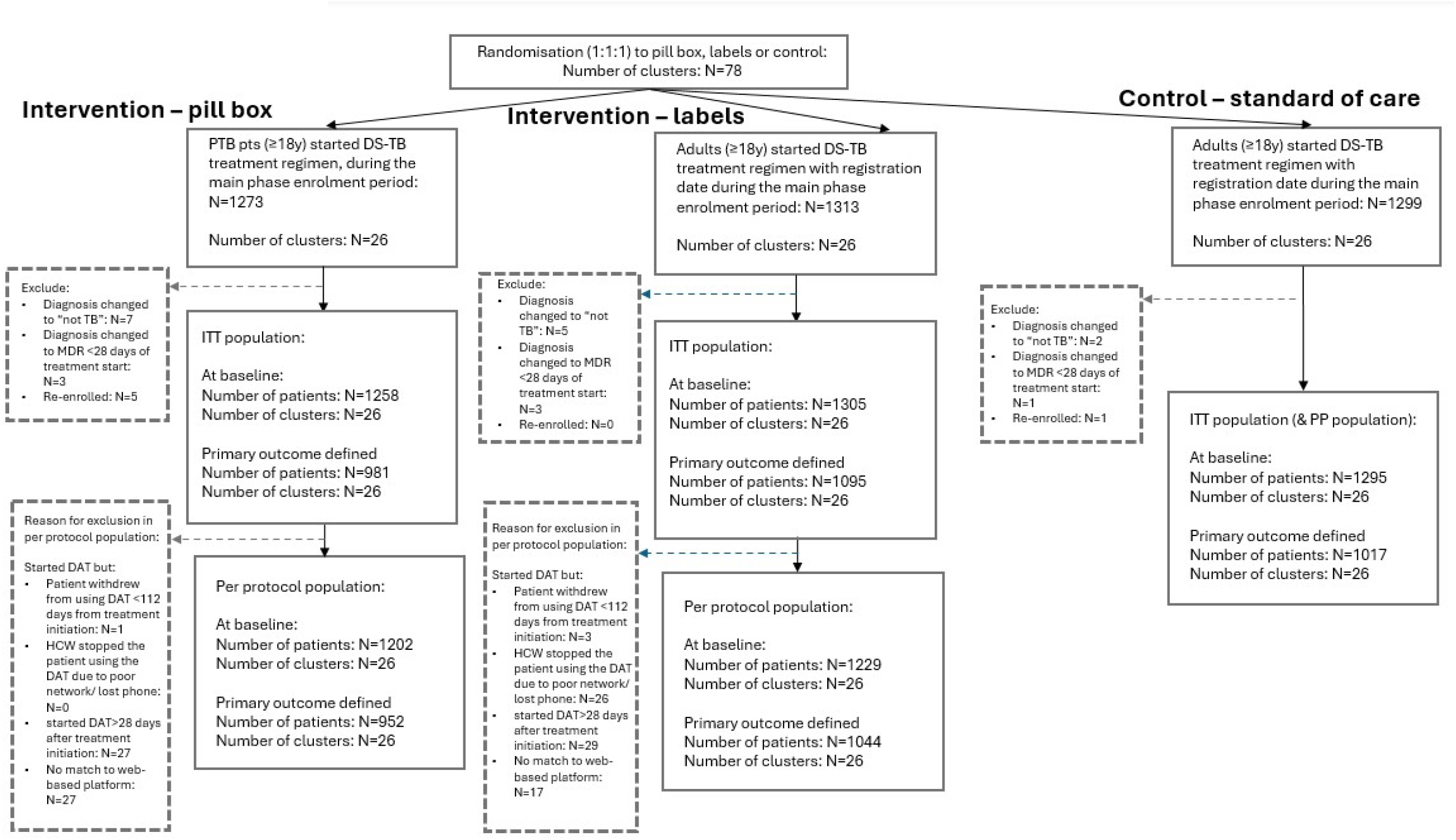
CONSORT diagram.

The demographic, social, and clinical characteristics of the participants are shown in Table 1. Of the 78 clusters, 36 were in Addis Ababa and 42 were in Oromia. Additionally, five clusters were in rural areas, and five were in hospital settings. Overall, 40.6% of participants were female, the median age was 30 years (interquartile range [IQR] 24–40 years), 64.8% had bacteriologically confirmed pulmonary TB, 6.6% had a previous history of TB, 12.6% were living with HIV, 49.3% had at least primary education or above, and 46.9% were married/ cohabited. There was a slight imbalance in age, sex, type of TB, and history of previous TB between study groups at baseline.

**Table 1:**
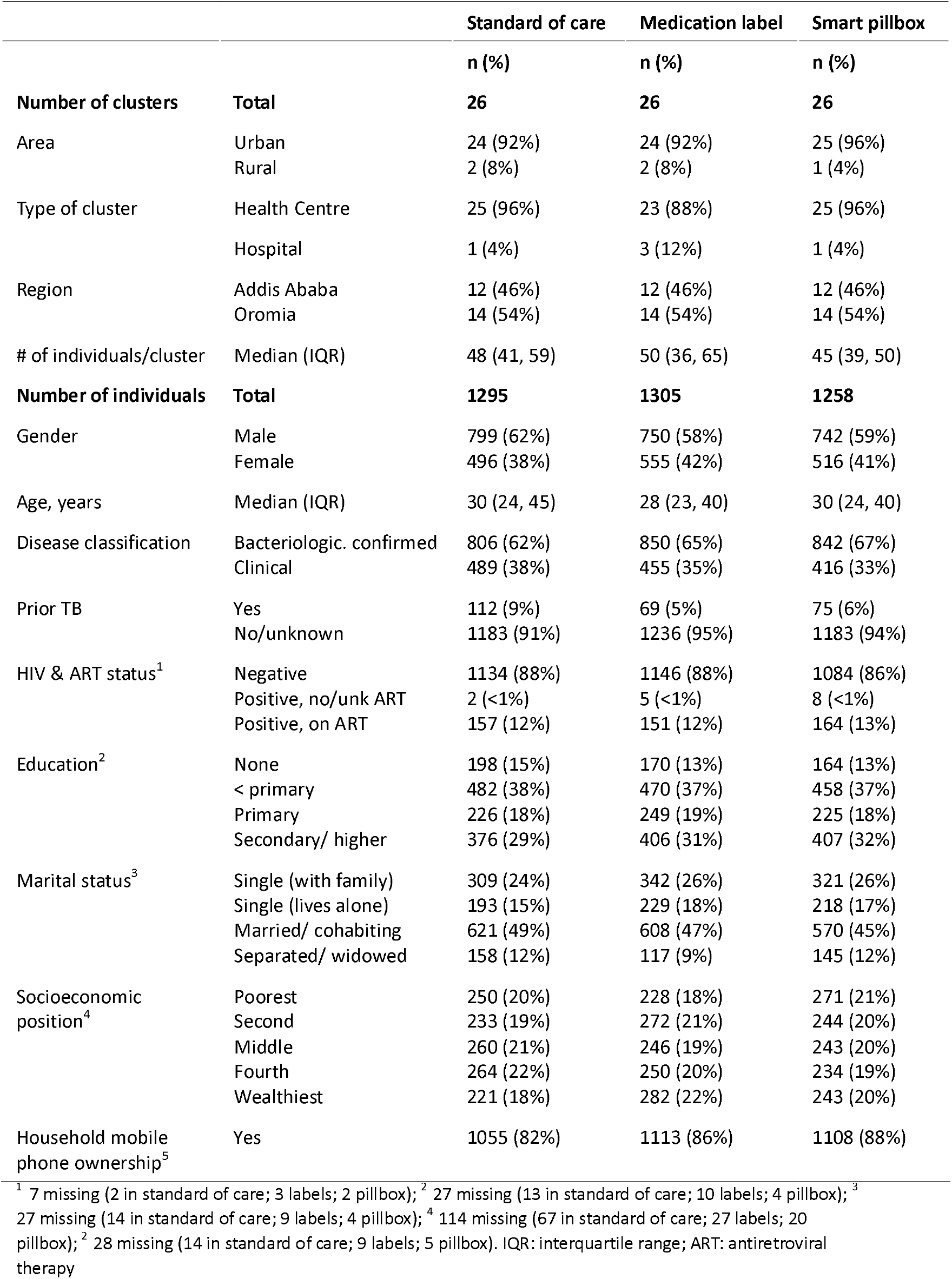
Baseline characteristics.

In the ITT population, the primary composite outcome data at 12 months were missing for 278 (21.5%) of 1295 participants in the standard of care, 210 (16.1%) of 1305 in the labels, and 277 (22.0%) of 1258 in the pillbox group (Figure 1). The main reason (99%) for missing outcome at 12 months was due to participants being unreachable by the research team. Using multiple imputation for missing outcomes, 95 (7.3%) of 1295 in the standard of care, 101 (7.7%) of 1305 in the labels and 93 (7.4%) of 1258 in the pillbox arm, satisfied the composite unfavourable outcome, based on the arithmetic mean of events from 25 imputations. Compared to the standard of care, there was no evidence of an effect in either the label (adjusted OR 1.14, 95% CI: 0.83 to 1.61; adjusted risk difference, 0.4 percentage points, 95% CI -1.8 to 2.6) or the pillbox (adjusted OR 1.04, 95% CI: 0.74 to 1.45; adjusted risk difference, 1.0 percentage points, 95% CI -1.2 to 3.1) arms, after adjusting for randomisation strata and baseline imbalance for sex, age group, bacteriological/clinical diagnosis, and previous history of TB, and HIV status and socio-economic position, both associated with missing outcome (Table 2). Results were similar in unadjusted, complete case, and per-protocol analyses.

**Table 2:** Primary and secondary outcomes by study arm.

|  | SoC | Label | Pillbox | Label vs SoC |  | Pillbox vs SoC |  |
| --- | --- | --- | --- | --- | --- | --- | --- |
| Primary outcome | n/N(%) | n/N(%) | n/N(%) | AOR<sup>3</sup> 95% CI | P-value | AOR<sup>3</sup> 95% CI | P-value |
| Unfavourable outcome (ITT)<sup>1</sup> | 95<sup>4</sup>/1295(7.3%) | 101<sup>4</sup>/1305(7.7%) | 93<sup>4</sup>/1258(7.4%) | 1.14 (0.83-1.61) | 0.40 | 1.04 (0.74-1.45) | 0.84 |
| Unfavourable outcome (ITT)<sup>2</sup> | 86/1017(8.5%) | 95/1095(8.7%) | 83/981(8.5%) | 1.09 (0.77-1.54) | 0.62 | 0.99 (0.69-1.41) | 0.95 |
| Unfavourable outcome (PP)<sup>2</sup> | 86/1015(8.5%) | 81/1043(7.8%) | 80/951(8.4%) | 0.94 (0.66-1.35) | 0.73 | 0.98 (0.68-1.40) | 0.9 |
| **Secondary outcome** |  |  |  |  |  |  |  |
| Poor treatment outcome (ITT)<sup>2</sup> | 65/1288(5.1%) | 70/1303(5.4%) | 59/1253(4.7%) | 1.15 (0.81-1.64) | 0.44 | 0.98 (0.68-1.42) | 0.92 |
| Lost to follow-up (ITT)<sup>2</sup> | 21/1288(1.6%) | 8/1303(0.6%) | 13/1253(1.0%) | 0.37 (0.15-0.95) | 0.039 | 0.67 (0.29-1.52) | 0.337 |

SOC: standard of care; ITT: Intent-to-Treat population; PP: Per-Protocol population; AOR: Adjusted Odds Ratio; CI: confidence interval.
<sup>1</sup> based on multiple imputation for those with missing outcomes; <sup>2</sup> complete case analysis; <sup>3</sup> analysis using imputation adjusted for randomization strata and baseline imbalance (sex, age groups, disease classification, previous TB), as well as HIV status, and socio-economic position and complete case analyses adjusted for randomization strata and baseline imbalance (sex, age groups, disease classification and previous TB), both taking into account clustering; <sup>4</sup> arithmetic mean of the number of events across the 25 imputations.

Of the 264 events satisfying the composite unfavourable outcome, the majority were either death (49.6%; 131/264), loss to follow-up on treatment (15.9%; 42/264), or TB recurrence at 12 months (26.5%; 70/264).

For the secondary outcome analysis, nearly 95% of participants in the ITT had either cure or treatment completion as their end-of-treatment outcome across the three study groups. Compared to the standard of care, there was no evidence of an effect of pillbox intervention on either the poor end-of-treatment outcome (adjusted OR 0.98, 95% CI: 0.68 to 1.42) or loss to follow-up (adjusted OR 0.67, 95% CI: 0.29 to 1.52). Similarly, there was no evidence of the effect of label intervention on the poor end-of-treatment outcome (adjusted OR 1.15, 95% CI: 0.81 to 1.64), but there was weak evidence of a reduction of loss to follow-up (adjusted OR 0.37, 95% CI: 0.15 to 0.95) in the label arm.

The prespecified subgroup analysis revealed no differences in the intervention effects on the primary composite outcome among subgroups based on participants’ age, sex, HIV status, type of TB, socio-economic position, place of residence, region, or strata. Sensitivity analyses conducted for the primary composite outcome and secondary outcomes aligned with the findings from the primary analyses. The arithmetic means of digital confirmation of doses taken was 89.8% and 79.9% in the pillbox and label arms, respectively. The intra-cluster correlation coefficients adjusting for randomisation strata were 0.05, 0.002 and 0.009 in the standard of care, labels and pillbox arm, respectively.

## Discussion

This is the first large-scale three-arm cluster-randomized trial evaluating the effectiveness of two types of DATs in the African region. It is also one of the few trials, globally, measuring the impact of DATs on TB recurrence. A total of 3858 participants with pulmonary DS-TB were enrolled by TB focal staff across 78 health facilities within two regions of Ethiopia. Neither DAT intervention had an effect on the primary composite unfavourable outcome, in both the ITT and PP populations, though there was a weak signal that labels reduced loss to follow-up. Among those with a treatment outcome of being cured or completed treatment at treatment completion, recurrence after 12 months from treatment initiation was 2.4%. Pre-specified subgroup analyses did not yield any effects on the primary outcome.

While other studies show mixed results on the effectiveness of DATs (12), our study concurs with findings from recent large cluster-randomized trials, indicating no effect on treatment outcomes and recurrence (13-15). These findings carry important implications for TB management strategies, suggesting that while DATs may offer potential benefits in improving treatment adherence, they may not translate into tangible improvements in treatment success rates or reduction in recurrence. This challenges the assumption that improving adherence alone can substantially impact TB outcomes and underscores the need for comprehensive interventions addressing other factors influencing treatment outcomes.

In our study, among participants who were cured/completed treatment, TB recurrence, measured by a single culture at 12 months, was 2.4% (70/2899) for those initially bacteriologically confirmed or restarting treatment at 12 months. This compares to the China (13) and South Africa (14) studies which found 2% (42/2105) and 3.8% (84/2190) recurrence, respectively, both measured over 12 months from the end of treatment. TB recurrence offers a comprehensive assessment of treatment efficacy beyond programmatic endpoints such as treatment completion or cure. TB treatment outcomes, which particularly rely on treatment completion, are problematic, as they reflect an insensitive measure of treatment success. This approach primarily measures whether participants adhere to their prescribed TB treatment within a set timeframe, overlooking long-term outcomes of TB recurrence. However, in the context of large pragmatic studies, this would entail sputum sampling for TB, sufficient sample size, long-term follow-up after the end of treatment, and robust surveillance systems, which are logistically demanding and costly.

Although the interventions did not influence poor end-of-treatment outcomes, there was weak evidence suggesting a reduction in loss to follow-up for the label intervention. Interestingly, this was not shown in the pillbox arm where an active daily medication reminder was provided. This observation warrants careful interpretation, given the small number of events (21 in standard of care; 8 in labels; 13 in pillbox), and 52 clusters reporting no losses to follow-up (16/26 standard of care; 20/26 in labels; 16/26 pillbox). Phone ownership varied by study arm, with a lower percentage in the standard of care, potentially resulting in death being incorrectly recorded as loss to follow-up in this arm. Further, there was one cluster in the standard of care arm whose catchment area included a shelter for the vulnerable, where 8.1% (5/62) of study participants were lost to follow-up, much higher than other clusters. A post-hoc analysis that excluded this facility revealed no effect of the labels on reducing loss to follow-up (adjusted OR 0.46, 95% CI: 0.18 to 1.18). Of all unfavourable outcomes, death on treatment accounted for the majority of events, with 47% (40/86) in the standard of care, 55% (52/95) in labels, and 47% ([39/83) in pillbox arm. Given the high mortality observed in the first two months (30), it is unlikely that the DAT interventions alone could influence the end-of-treatment outcome.

On the contrary, two individually randomised trials of a DAT intervention combined with other non-digital components, have demonstrated improved treatment outcomes. The intervention group in the Keheala study (25), conducted in Kenya, received a multicomponent intervention using daily text dosing reminders, motivational messages, adherence follow-up by a research team, and interaction with a support team. This intervention improved treatment success primarily by reducing loss to follow-up. Recent findings from a trial in Tibet (16), conducted in a very rural setting, where 25% of participants had microbiologically confirmed TB and the median age of 56 years, reported improved treatment outcomes among participants in the intervention arm versus standard of care. The intervention included a comprehensive package of interventions using medication monitors with voice reminders, health education and personalised support to use the DAT. The improvement in treatment outcomes again was primarily through reducing loss to follow-up. Of note, only 73% of trial participants achieved favourable outcomes in the standard of care arm. Other studies have shown improved treatment outcomes, particularly within specific populations (21, 24). In Uganda, the intervention group received daily dosing reminders and a weekly automated interactive voice response check-in, resulting in a reduced loss to follow-up for 52% of the per-protocol population compared to the standard of care phase (24). Similarly, a trial in Peru utilized a real-time medication event reminder monitor with additional assistance on its use by a treatment supporter, resulting in improved treatment outcomes (21). None of these trials measured TB recurrence, though passive follow-up is planned for the Tibet study. Consequently, the long-term effectiveness of these interventions remains unknown.

Our findings align closely with recent large cluster-randomized trials of pillbox interventions from China and South Africa (13, 14). Similar to our study, these trials included tuberculosis recurrence in their composite unfavourable outcome and did not find evidence of the pillbox intervention impacting on composite unfavourable outcomes (13, 14). The geometric mean for TB treatment adherence, measured by pillbox openings, was 89% in these trials, higher compared with standard of care participants who used the pillbox in silent mode. This, however, did not translate into improved health outcomes. In our trial, adherence was not measured in the standard of care arm, but real-time digital confirmation was 89.7% in the pillbox and 79.9% in the label groups. This indicates a potential opportunity for healthcare providers to offer additional adherence support to individuals experiencing adherence challenges. A recent systematic review suggests that the effectiveness of DATs depends on the DAT type and the implementation setting (12). Video-Supported Therapy interventions were found to be beneficial in high-income settings, but SMS-based and pillbox interventions were not consistently impactful in improving TB treatment outcomes in low- and middle-income countries, suggesting a need for further refinement in the design and delivery approaches (12).

There are limitations to this study. The trial was not masked and so may be more susceptible to performance bias, as participants and TB focal staff across all arms may behave differently. We had no measurement of adherence in the standard of care, which prevented us from evaluating the impact of study arms on adherence. Our study was adequately powered to detect a one-third reduction in the primary composite outcome, considering a baseline unfavourable outcome rate of 17-20% in the standard of care. These assumptions were based on pre-trial treatment outcome data that were not consistent with the standard of care poor outcome realised in the trial; a lower percentage was observed (8.5%). However, unfavourable outcome percentages were similar by arm, suggesting we did not miss an impact of the intervention due to reduced power. The lower percentage of unfavourable outcomes compared with pre-trial data may be attributed to a uniform reduction in all three arms due to study effects, similar to Hawthorne effect, or higher percentage of bacteriologically confirmed TB at the initiation of treatment (two-thirds of the trial population), aided by GeneXpert and sputum smear, which in turn could lead to improved health outcomes. The determination of TB-free status at the 12-month follow-up for those clinically diagnosed with TB relied on passive detection through self-report of no initiation of TB treatment, rather than other diagnostic approaches, which may impact the accuracy of detecting TB recurrence. One-third of the recurrences were based on being culture-positive for *M. tuberculosis*. We did not abstract additional data from the TB register for participants who reported starting TB treatment at 12 months, some of which would have been bacteriologically confirmed. The trial was conducted during the COVID-19 pandemic which has altered the standard of care and influenced the delivery of TB care. Less frequent patient visits to the health facilities occurred and despite this, outcomes in the standard of care arm were similar to the intervention. The trial’s strength lies in its pragmatic nature, large sample size, and wide implementation across diverse health facilities, delivered by TB focal staff instead of research personnel. Additionally, it included a 6-month follow-up post-treatment to evaluate TB recurrence, making it one of the few TB DAT trials. High feasibility, acceptability and uptake of the interventions were observed in the study setting.

In conclusion, this is the largest pragmatic DAT trial in sub-Saharan Africa in which we evaluated the effectiveness of two commonly used DATs in reducing unfavourable treatment outcomes including TB recurrence in persons with pulmonary DS TB. The DAT interventions did not impact unfavourable treatment outcomes, except for a reduction in loss to follow-up in the labels arm, though with weak evidence. Subgroup analyses revealed consistent findings across demographic and clinical characteristics, with sensitivity analyses supporting the robustness of the primary result. These findings underscore the need for further exploration of integrating DATs with non-DAT interventions to optimize their effectiveness and achieve positive health outcomes in TB management strategies. Additionally, our study highlights the challenge of improving TB outcomes solely through adherence interventions and emphasizes the complexity of factors contributing to treatment success. Moving forward, further research and innovation are needed to effectively address the challenges posed by TB recurrence and improve treatment outcomes.

## Data Availability

The deidentified individual participant data underlying the results in this article, along with the study protocol and statistical analysis plan will be accessible on the London School of Hygiene & Tropical Medicine Data Compass (https://datacompass.lshtm.ac.uk/). These will be available without restrictions after October 1st, 2025.

## Contributors

The study’s concept and design were developed by AWT, NF, CFM, GTW, ZM, JvR, KvK, AB, TL, DJ and KLF. Data collection was handled by AWT, MS, TA, ZM, MB, AS, DA, DG, HY, NY, ZS and NM. Data analysis was conducted by AWT and KLF. AWT and KLF drafted the manuscript. All authors were involved in interpreting the data and approving the final version submitted for publication. The corresponding author confirms that all listed authors meet the criteria for authorship. Each author had full access to the data in this study and bore the final responsibility for submitting the manuscript for publication. AWT and KLF accessed and verified the underlying data presented in this study.

## Data sharing

The deidentified individual participant data underlying the results in this article, along with the study protocol and statistical analysis plan will be accessible on the London School of Hygiene & Tropical Medicine Data Compass (https://datacompass.lshtm.ac.uk/). These will be available without restrictions after October 1^st^, 2025.

## Acknowledgements

This study was supported by Unitaid (2019–33-ASCENT) through the Adherence Support Coalition to End TB (ASCENT) project (https://www.digitaladherence.org/). Technical Advisory Group: Professor Achilles Katamba (chair) Makerere University, Uganda; Professor Dr Mary Ann Lansang, University of the Philippines, Manila, Philippines; Professor Tobias Chirwa, University of the Witswatersrand, South Africa; Dr Getnet Yimer, Centre for Global Genomics & Health Equity, Department of Genetics, Perelman School of Medicine, University of Pennsylvania, Philadelphia, Pennsylvania, USA; Dr Natalia Lytvynenko, National Research Institute of TB and Lung Diseases, Ukraine; Dr Andrew Martin Kilale, National Institute for Medical Research, Muhimbili Medical Research Centre. Tanzania. We also wish to express our gratitude to the participants who consented to be part of this study, the National Tuberculosis Program at the Ministry of Health in Ethiopia, the Disease Prevention and Control team at the regional health bureaus in Addis Ababa city and Oromia regional state, as well as the zonal, town, sub-city, and woreda health offices, and the TB focal points at each facility for their support in implementing the ASCENT project.

